# Evaluating the Utility of EZC Pak, a 5-Day Combination Echinacea-Zinc-Vitamin C Dose Pack with or without Vitamin D, in the Management of Outpatient Upper Respiratory Infections

**DOI:** 10.1101/2022.10.18.22280622

**Authors:** Aditya Radhakrishnan, Stephanie Spencer, Naveena Yanamala, Sarath Malepati

## Abstract

**Background:** Growing antibiotic resistance is among the most serious threats to healthcare systems and public health globally with antibiotic misuse considered a leading driver of this problem. One of the largest areas of antibiotic misuse is in outpatient upper respiratory infections (URIs), the most common infection in humans. The purpose of this research is to evaluate the efficacy of EZC Pak, a combination Echinacea-Zinc-Vitamin C dose pack with or without Vitamin D, on the duration of illness and symptom severity of non-specific URIs as an alternative to antibiotics when none are deemed clinically necessary. A secondary analysis was carried out on patient satisfaction with using EZC Pak.

**Methods:** A total of 360 patients across the United States were enrolled and randomized in a double-blind manner across two intervention groups, EZC Pak, EZC Pak+Vitamin D, and one placebo group. The study was conducted virtually utilizing a smartphone-based app to screen, enroll and capture study data of the participants. Once a study participant reported the first symptoms of a URI, they were advised to take the intervention as directed and complete the daily symptom survey score until their symptoms resolved.

**Results:** The average EZC Pak participant recovered 1.39 days faster than placebo (*p* = 0.017) than the average placebo participant. The average EZC Pak participant reported a 17.43% lower symptom severity score versus placebo (*p* = 0.029). EZC Pak users reported 2.9 times higher patient satisfaction versus users of the placebo (*p* = 0.012). The addition of Vitamin D during this acute phase of illness neither benefited nor harmed illness duration or symptom severity.

**Conclusions:** The findings support the potential use of EZC Pak as a viable alternative to patient request for antibiotics when none are deemed clinically necessary at the time of initial clinical presentation. The decision to replete vitamin D in the acute phase of URI is an individualized decision left to the patient and their clinician. EZC Pak may play a critical role in improving outpatient URI management and antibiotic stewardship. (ClinicalTrials.gov number, <u>NCT04943575</u>.)

## Introduction

Growing antibiotic resistance and the concomitant rise of drug-resistant pathogens (superbugs) is among the most serious threats to healthcare systems and public health globally. A UK government-commissioned analysis on rising antimicrobial resistance suggests that current growth rates may lead to more deaths from superbugs than cancer by 2050 at a USD $100 trillion cost.^1^

While there are a host of factors contributing to this multifaceted, complex problem, the World Health Organization (WHO) considers the inappropriate use of antibiotics when none are necessary to be the leading driver of the growth of superbugs.^2^ The seriousness of this expanding crisis has reached a stage where the WHO is requesting action across government and the private sector to develop strategies that prioritize the reduction of antimicrobial resistance.^2,3^

While this problem is pervasive across the healthcare system, one of the biggest areas of antibiotic misuse is in the outpatient management of upper respiratory infections (URIs). URIs are the most common infection in humans. Eighty percent of upper respiratory infections are caused by viruses.^4^ Antibiotics only treat bacterial infections, not viral infections. Despite this, antibiotic usage without evidence of a bacterial infection remains a critical problem. This problem has been highlighted most recently in the surge of antibiotic overuse globally during the COVID-19 pandemic.^5,6^

While vaccines and antiviral medications exist, wider adoption remains limited due to low access or low uptake of such interventions in certain geographies and communities, high viral mutation rates rendering treatments potentially ineffective^6,7^, and weighing the efficacy and risk benefit ratio of using costly interventions for what amount to in most cases, mild and self-limited disease.^8^

Vitamins, minerals, and herbs to support the immune system’s clearance of infections have broadly shown mixed benefits. This has been in part due to the lack of uniform standards in preparation, formulation, potency, and actual usage.^9^ The dosages necessary to confer therapeutic benefit demonstrated in supportive studies are often much higher than the dosages found in standard products commonly found over the counter in pharmacies and drug stores. A key advantage of the potential use of vitamin, mineral, and herbal preparations as a tool in URI management is the reduced risk of antimicrobial resistance, reduced exposure to the potential side effects of drugs, and the long-term preservation of antibiotic and antimicrobial efficacy when they are critically needed.

In the case of the Western herb *Echinacea*, the strongest data for its potential use in the treatment of URIs is likely in the form of *Echinacea purpurea*.^10,11,12,13,14,15,16^ In the case of the mineral zinc, the greatest potential benefit of its usage in the management of URIs may be in ionizable forms of zinc, such as zinc acetate.^17,18,19,20^ The utility of vitamin C supplementation in either the prevention or treatment of URIs has yielded mixed results in randomized clinical trials (RCTs), but may provide more benefit in the prevention of URIs in patients doing heavy exercise and undergoing similar short term physical stress.^21,22,23^

There has been increased interest in the potential role of vitamin D in URI management during the COVID-19 pandemic. While data is mixed^24,25^, some current data shows there may be potential benefit in vitamin D supplementation in reducing the incidence of URIs.^26,27,28^ However, a study evaluating early, acute repletion of vitamin D with high enteral bolus during active COVID-19 illness in intensive care unit patients did not show benefit in reducing 90-day all-cause mortality.^29^

While there are a number of studies evaluating the use of individual vitamins, minerals, and herbs in the management of URIs, RCTs evaluating the potential role of combinations of herbs, vitamins, and minerals that individually have demonstrated data supporting their individual usage remain limited. The author’s interest in examining the potential use of such treatment modalities stems from the WHO’s public request and is derived from a desire to equip clinicians and patients with tools that reduce inappropriate antibiotic use during viral URI. The end goal is to reduce the potential risk of the induction and spread of antibiotic resistance.

The purpose of this specific study is to evaluate the feasibility of using EZC Pak, a 5-day dose pack of Echinacea purpurea, zinc acetate, and vitamin C to reduce the duration of illness and symptom severity in non-specific upper respiratory infection. A secondary endpoint was evaluating patient satisfaction with receipt of the intervention. Given both public and academic interest in a potential role for vitamin D in URI management, a second intervention arm adding vitamin D to the base dose pack of Echinacea purpurea, zinc, and vitamin C was carried out.

## Methods

Institutional review board (IRB) approval of the study protocol was carried out by Argus IRB. A total of 360 individuals, male or female over 18 years old, were then enrolled by a third-party clinical research organization (CRO), to participate in the study. The CRO utilized an algorithm to randomize the patients into three different arms with comparable demographics – placebo, EZC Pak, or EZC Pak + Vitamin D (EZC Pak+D) in a double blinded manner. The placebo was composed of rice concentrate.

A total of 180 individuals were enrolled in the placebo control arm. A total of 120 individuals were enrolled in the EZC Pak intervention arm. A total of 60 individuals were enrolled in the EZC Pak+D intervention arm. Enrollment was completed during the initial 90 days and the intervention component was carried out over the subsequent six months.

Individuals with the following medical conditions were excluded: ragweed allergy, chronic seasonal allergies, liver disease, autoimmune or connective tissue disorder (e.g., rheumatoid arthritis, lupus, multiples sclerosis, HIV), alcohol consumption more than 7 drinks per week, or more than 3 drinks per occasion, routine recreational drug use, renal disease, and females that were pregnant, wanted to become pregnant for the duration of the study, or who were breastfeeding.

The study was conducted virtually with the CRO’s technology platform utilized to screen, enroll and capture study data of the participants. Study enrollees had to actively participate in the study intervention only when they had a URI. Once participants had a URI, they took the double blinded test product as directed and completed the daily symptom survey score until their symptoms resolved.

Participants also recorded any adverse or ill effects any time after taking the test product and for any final adverse events upon completion of the exit form. Participants also recorded if they took any additional medications during the course of their URI.

Once a study participant reported the first symptoms of a URI, they were advised to take the intervention (i.e., placebo, EZC Pak, or EZC Pak+D) taper dosed over a five-day period. The participant was instructed to take the intervention 4 times a day (every 6 hours) on Day 1, 3 times a day (every 8 hours) on Day 2, and 2 times a day (every 12 hours) on Day 3 through Day 5 (Figure 1).

**Figure 1.**
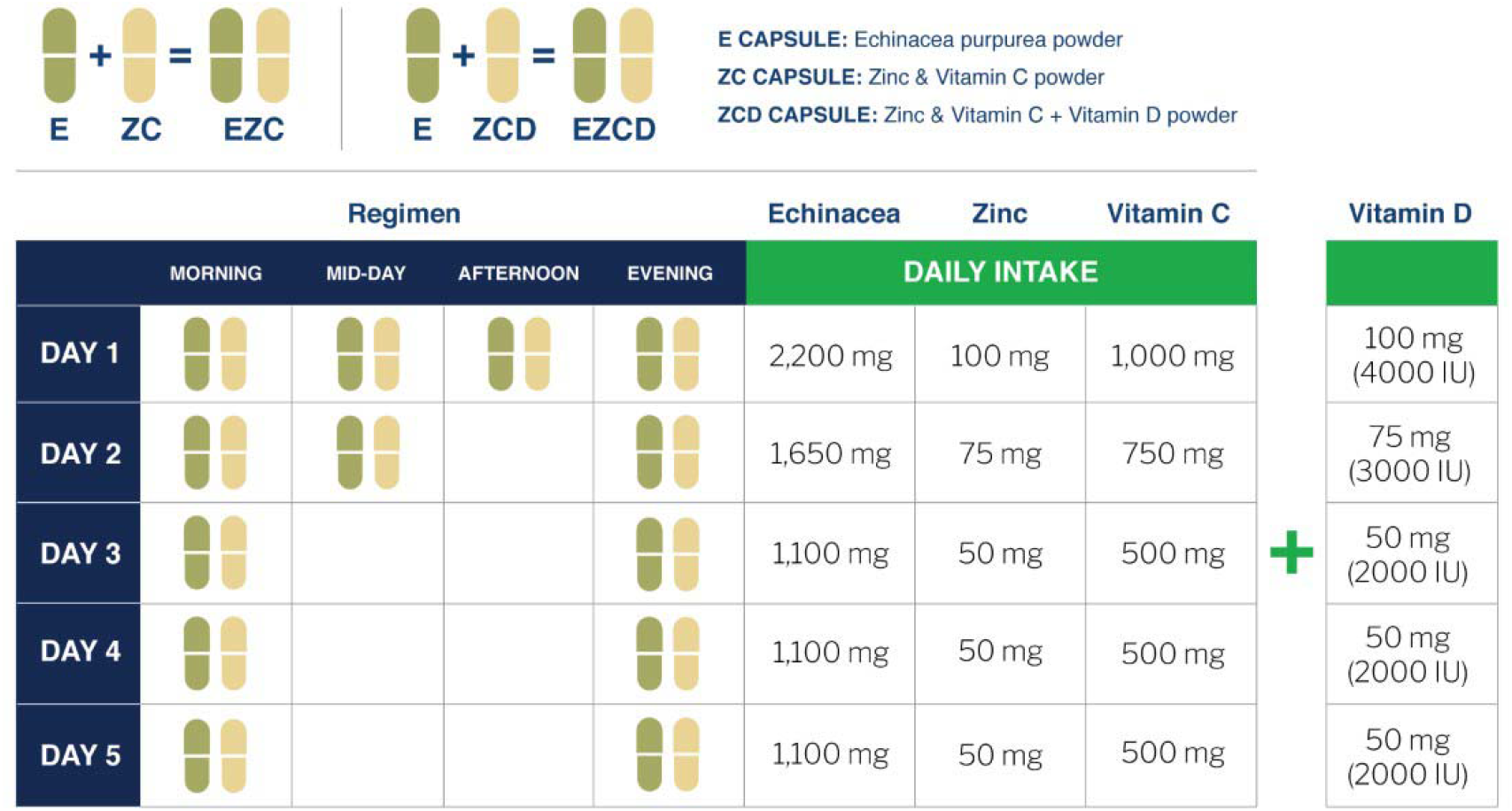
Visual representation of the dosing scheme over the five-day treatment period.

### Analysis

In order to evaluate the performance of the intervention, a comparative analysis on the illness of trial participants from the intervention arms EZC Pak and EZC Pak+D was performed versus the placebo control arm. There were 360 subjects enrolled in this study, well distributed across the mainland US (Figure 2). The age range across the sample was between 22 years and 88 years old. The average age for the placebo group was 57 years old, the EZC group was 55 years old, and the EZC+D group was 54 years old. Most participants were White or Caucasian. The other ethnicities represented less than 10% of the sample within each intervention arm (Table 1).

**Figure 2.**
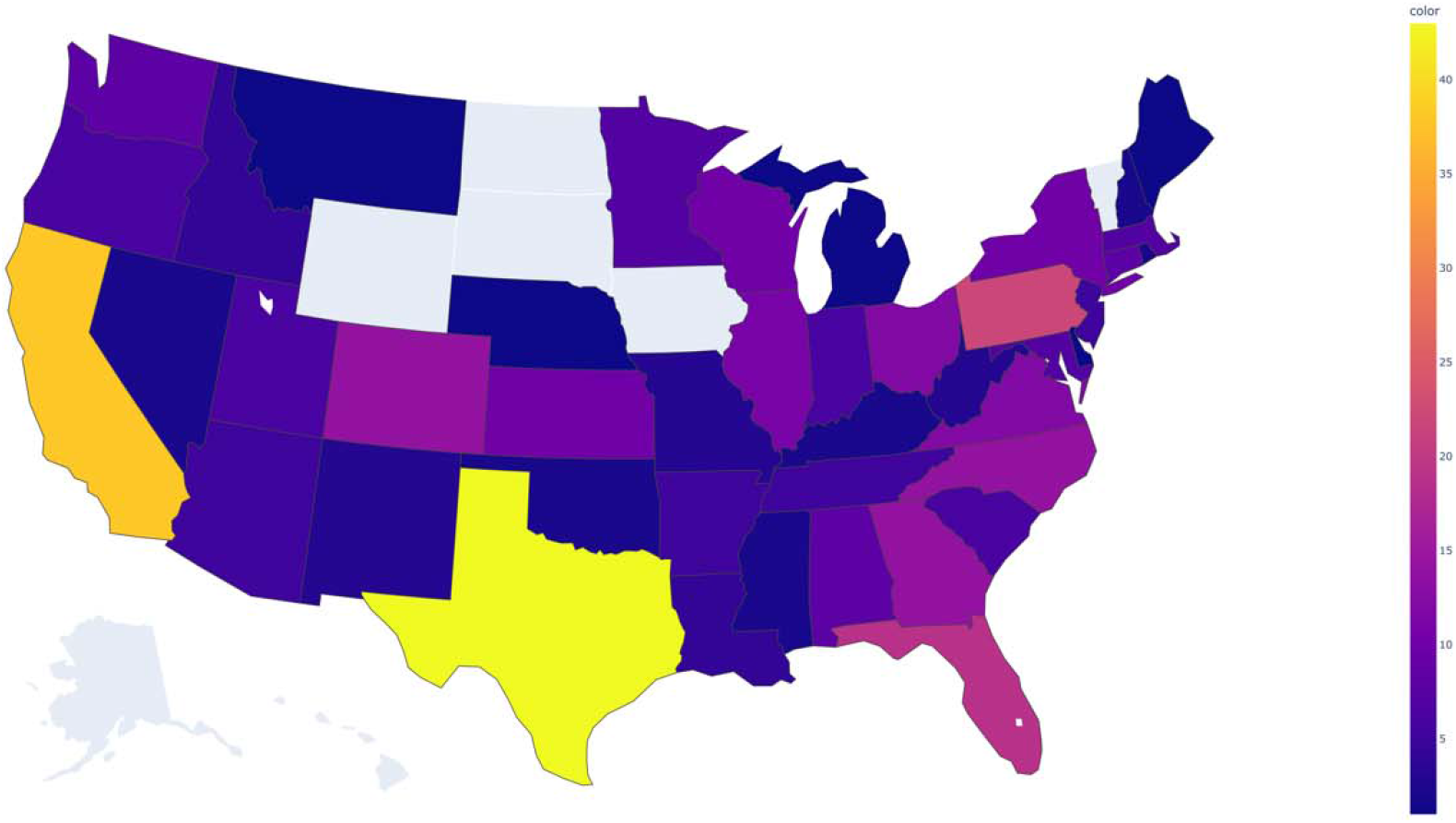
Choropleth Map of Geographic Distribution of Study Participants

**Table 1.** Demographic Characteristics of Participants

| Variable | Placebo Group<br>(n = 165) | EZC Group<br>(n = 123) | EZC D Group<br>(n = 72) |
| --- | --- | --- | --- |
| **Gender n (%)** |  |  |  |
| Male | 44 (26.7) | 28 (22.8) | 19 (26.4) |
| Female | 121 (73.3) | 95 (77.2) | 53 (73.6) |
| **Age (years)** |  |  |  |
| Mean ± SD | 56.9 ± 13.32 | 54.5 ± 14.83 | 53.8 ± 14.58 |
| Median | 59 | 58 | 56 |
| Min, Max | 22, 86 | 22, 88 | 23, 78 |
| **Ethnicity n (%)** |  |  |  |
| White or Caucasian | 117 (70.9) | 100 (81.3) | 60 (83.3) |
| Black or African American | 12 (7.3) | 6 (4.9) | 4 (5.6) |
| Hispanic or Latino | 14 (8.5) | 7 (5.7) | 4 (5.6) |
| East Asian | 6 (3.6) | 1 (0.8) | 1 (1.4) |
| South Asian/Indian subcontinent | 6 (3.6) | 1 (0.8) | 1 (1.4) |
| Native American or Alaska Native | 2 (1.2) | 1 (0.8) | 1 (1.4) |
| Middle Eastern or North African | 0 | 1 (0.8) | 0 |
| Multiracial or Multiethnic | 7 (4.2) | 4 (3.3) | 1 (1.4) |
| Other | 1 (0.6) | 2 (1.6) | 0 |
| **Household income n (%)** |  |  |  |
| \$30,000 or less | 53 (32.1) | 22 (17.9) | 12 (16.7) |
| \$30,000 to \$60,000 | 40 (24.2) | 35 (28.5) | 23 (31.9) |
| \$60,000 to \$100,000 | 31 (18.8) | 36 (29.3) | 20 (27.8) |
| \$100,000 to \$150,000 | 25 (15.2) | 19 (15.4) | 10 (13.9) |
| \$150,000 or more | 16 (9.7) | 11 (8.9) | 7 (9.7) |
| **Education level n (%)** |  |  |  |
| No degree | 4 (2.4) | 0 | 1 (1.4) |
| High school or equivalent | 29 (17.6) | 27 (22.0) | 15 (20.8) |
| Bachelor's degree | 67 (40.6) | 49 (39.8) | 28 (38.9) |
| Master's degree | 43 (26.1) | 28 (22.8) | 13 (18.1) |
| PhD | 1 (0.6) | 3 (2.4) | 3 (4.2) |
| Other | 21 (12.7) | 16 (13.0) | 12 (16.7) |

Two evaluations were carried out. The primary evaluation assessed illness on the basis of daily symptom and vital sign reporting. Secondarily, we performed an analysis on subjective patient satisfaction. In addition to separate comparisons, EZC Pak and EZC Pak+D arms were also combined to include all participants from both pools.

Given that the entire adult population of the United States, or 258.3 million adults^29^, can be infected by URIs, a 90% confidence interval with a 5% margin of error was used to select a 360 patient sample size for this initial feasibility study.

We evaluated the performance of each illness via two metrics: Days To Recovery (DTR) and Symptom Severity During Recovery (SSDR). Two separate, parallel analyses were carried out for each to determine the efficacy of the intervention versus the placebo.

### Days To Recovery (DTR)

DTR is a metric that measures the total number of days in which a patient is experiencing symptoms of an illness while taking the intervention or placebo. DTR also directly lends itself for use in a log-rank analysis, which is used to compute statistical significance.

The primary limitation of DTR arises from the heterogeneous nature of the illness and the patient’s unique recovery. Despite aggressive daily oversight, some participants were lost in follow up, most particularly, but perhaps not surprisingly, in the placebo group (Table 2).

**Table 2.**
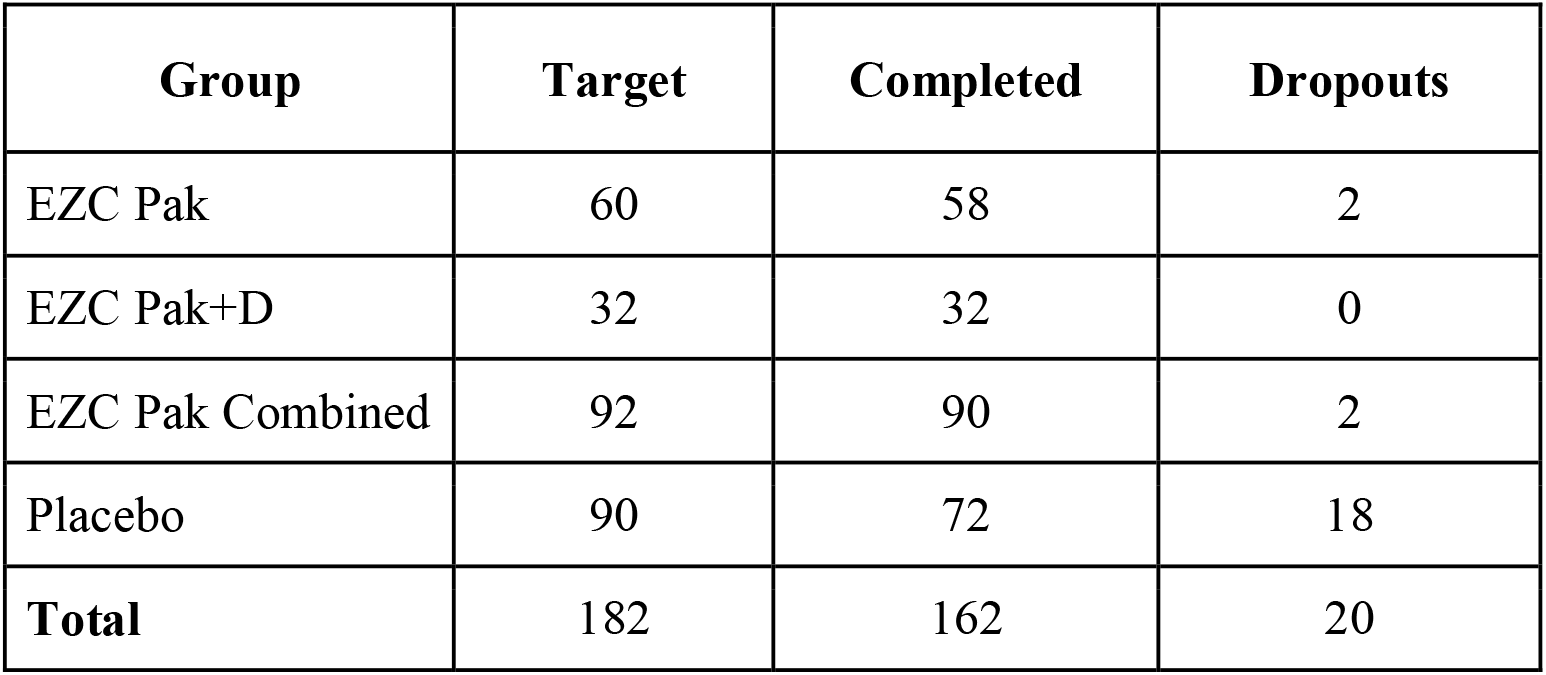
Distribution of Subject Completion and Dropout Status by Group

Interestingly, the 20% dropout rate in the placebo group was comparable to the proportion of patients, approximately 20%, one would expect to have a bacterial infection that may require antibiotics. In these instances of dropout, a DTR score of 14 days was imputed, equivalent to the average number of days a bacterial infection lasts if left untreated.

From the numerical results, the average EZC Pak participant recovers nearly one and a half days sooner than the average placebo participant (Table 3). From the KDE plot (Figure 3), we can similarly observe a higher density of early recoveries as compared to the placebo group, with EZC Pak group peaks coming slightly sooner. For EZC Pak and the EZC Pak Combined cohort, we see that this is a statistically significant improvement (*p* < 0.1). EZC Pak+D does not achieve the same level of statistical significance in the DTR analysis driven by the small sample size.

**Table 3.** Mean DTR Score and Improvement over Placebo with Log-Rank test

| Group | Mean DTR Score | Improvement Over Placebo | p-value (log-rank test; versus placebo) |
| --- | --- | --- | --- |
| EZC Pak | 5.82 days | 1.44 days | 0.022 |
| EZC Pak+D | 5.97 days | 1.29 days | 0.124 |
| EZC Pak Combined | 5.87 days | 1.39 days | 0.017 |
| Placebo | 7.26 days | N/A | N/A |

**Table 4.** Post-Filter SSDR Cohort

| Group | Original Size | Post-Filter Size |
| --- | --- | --- |
| EZC Pak | 58 | 40 |
| EZC Pak+D | 32 | 18 |
| EZC Combined | 90 | 58 |
| Placebo | 72 | 42 |
| All | 162 | 100 |

**Figure 3.**
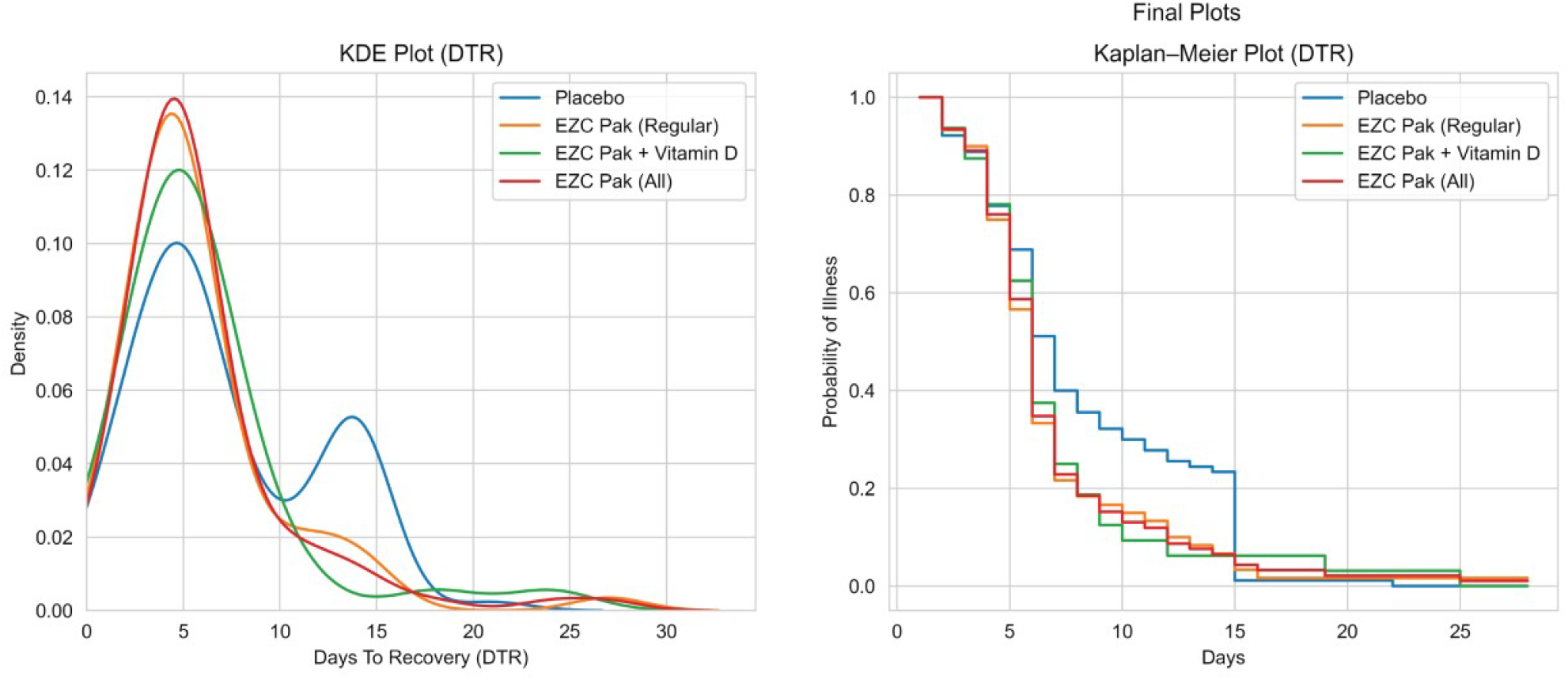
KDE and KM Plot for DTR Score.

### Symptom Severity During Recovery (SSDR)

SSDR is a metric that aims to capture information on how the symptoms of an illness progress during the recovery phase. Unlike DTR, it is intended to be disease agnostic, where scores range strictly from 0 to 1 regardless of the type and severity of an illness.

SSDR is motivated by the idea that the path to recovery provides useful insights into a participant’s disease response. In order to assess this, SSDR is fundamentally based on the area under an illness curve. Such illness curves are generated by measuring daily symptom magnitudes for each illness day, as counted for DTR.

Daily symptom magnitudes utilized the following questions:

1. Are you experiencing coughing?
2. Are you experiencing hoarseness?
3. Are you experiencing runny nose?
4. Are you experiencing nasal congestion?
5. Are you experiencing sneezing?
6. Are you experiencing scratchy or sore throat?
7. Are you experiencing headache?
8. Are you experiencing ear pain?
9. Are you experiencing fatigue?
10. Are you experiencing chills?
11. Are you experiencing shortness of breath or difficulty breathing?
12. Are you experiencing new loss of taste or smell?
13. Are you experiencing nausea?
14. Are you experiencing vomiting?
15. Are you experiencing diarrhea?
16. Do you have a fever?

Subjective responses for each of 15 symptoms were registered as either “None,” “Mild,” “Moderate,” or “Severe.” Each of these was mapped to a numerical score of 0, 1, 2, and 3 respectively. Additionally, along with the symptoms, the presence of a fever was included, where “No Fever,” “Between 99.9-100.4,” “Between 100.5-101.4,” “Between 101.5-103,” and “More than 103” was similarly mapped to 0, 1, 2, and 3, respectively.

The total “symptom severity” for a day was the sum of these numerical scores across all symptoms and temperature. Other vitals including respiratory rate, heart rate, blood pressure, and oxygen saturation were recorded in the study, but ultimately discarded due to the observed high unreliability of participant self-measurement and recording of these vital signs.

From these symptom scores, a sequence of daily symptom severities was generated, the sum of which is the area under the illness curve. In its raw form, the area represents the total discomfort or pain that a patient experiences during an illness. Reducing this is generally a foundational goal.

With respect to illness heterogeneity, simply taking the raw area under the illness curve brings the same challenges as DTR, only to a larger extent. As such, a scaling approach is used where the symptom severity is scaled according to the illness peak, and the timesteps are scaled based on the length of the recovery phase. This is also equivalent to the ratio of the observed area under recovery to the area under recovery if the illness had stayed static and not improved at all from the peak.

In effect, this results in a theoretical maximum SSDR of 1 and a minimum of 0. The lower the SSDR, the better. SSDR utilizes the area under the recovery phase and not the entire illness. To determine the start of recovery, the greatest one-day decrease in symptom severity was identified and then analyzed backwards until symptom severity stopped improving. In effect, this results in SSDR being a metric that focuses on the severity of symptoms.

SSDR can still be difficult to compute under certain circumstances. For example, in instances where a participant skipped multiple days of data entry, the exact shape of the illness curve becomes unknown. To the extent possible, this was addressed using a linear estimate of nearby datapoints, but such estimates become unreliable if an excess number of data entries are missing.

As such, some basic filters were utilized to assess subject participants with usable and ultimately clinically relevant SSDRs:

1. Participants with incomplete end of data entry reporting or more than 30% of their daily data entries missing
2. Illnesses that were less than three days or longer than ten days

This results in a SSDR cohort:

We observe a 15-20% improvement in SSDR when comparing the mean performance of the EZC Pak groups versus placebo (Table 5). In the KDE plot, we also observe the very clear leftward shift in EZC Pak group peaks (Figure 4). Most EZC Pak participants have notably lower SSDRs than their corresponding placebo counterparts.

**Table 5.**
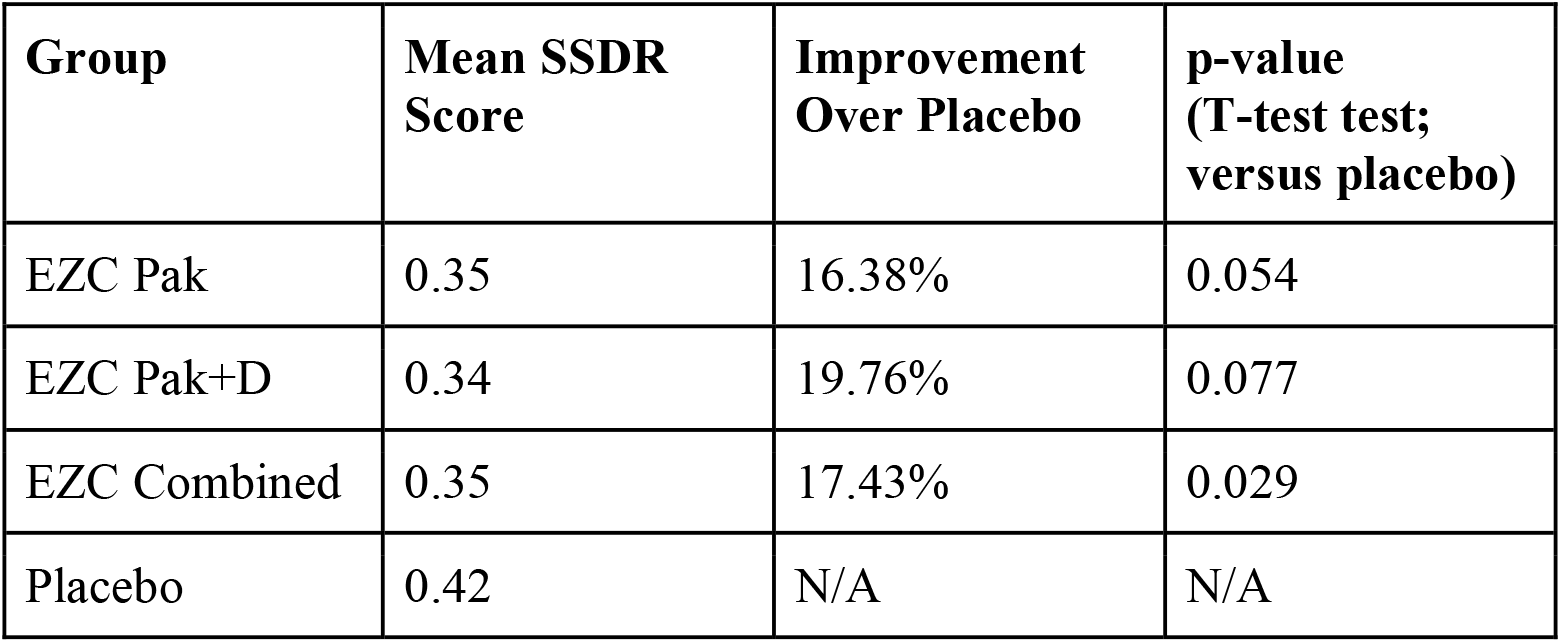
Mean SSDR Score and Improvement Over Placebo with t-test

**Figure 4.**
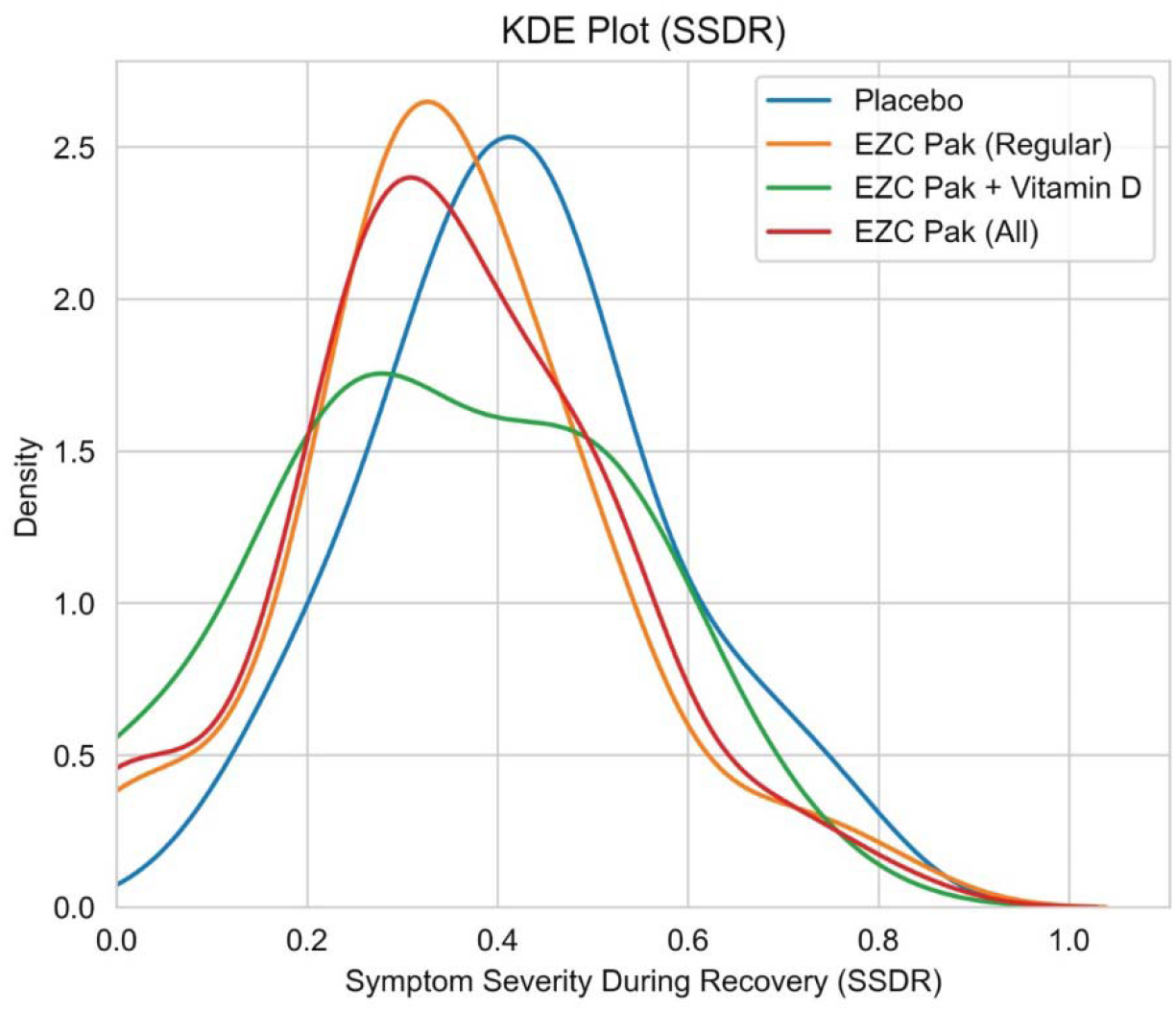
KDE Plot for SSDR Score

### Patient Satisfaction Survey Analysis

At the end of illness, participants were asked the following: “Do you believe the test product sped up your recovery time?” The options they could choose were “Yes,” “Maybe,” and “No.” These responses were mapped to scores 1, 0, and -1 respectively, enabling a scale that ranges [-1, 1]. Negative scores imply a lack of confidence in the product, zero implies ambivalence, and positive scores imply satisfaction. No filters were applied or imputations made.

All EZC Pak groups had a higher mean participant satisfaction than the placebo group, with the base EZC Pak performing approximately three times better than placebo (Table 6).

**Table 6.**
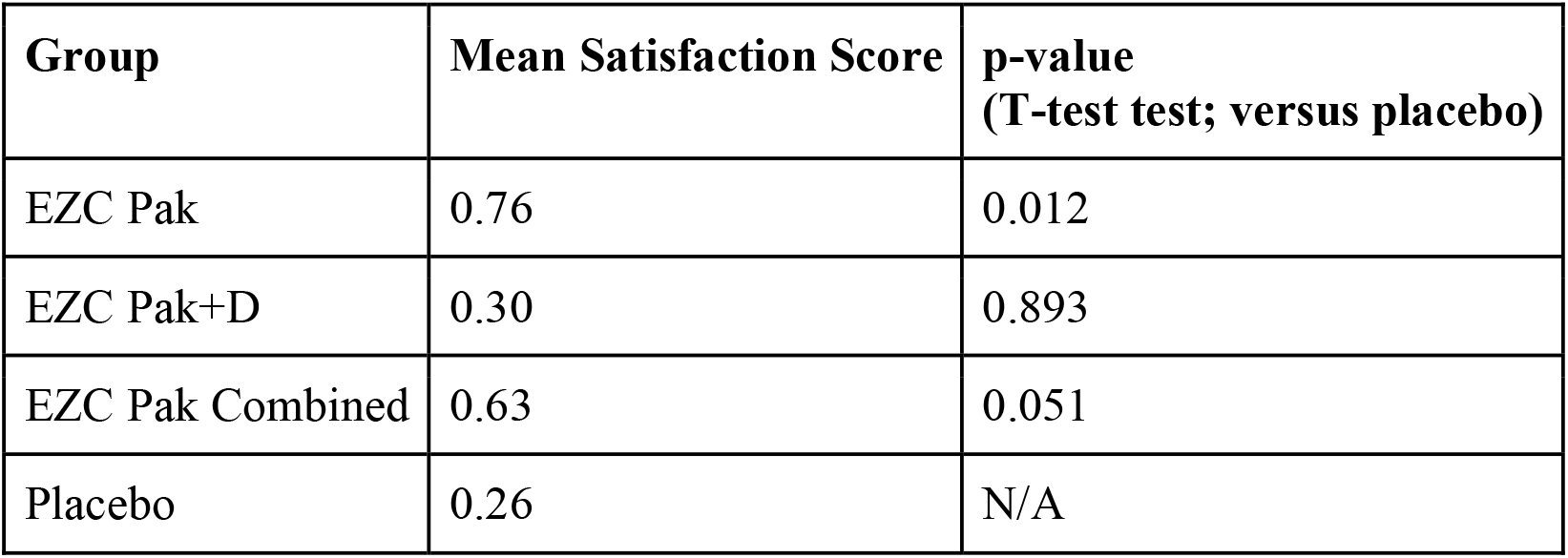
Mean Satisfaction Score with t-test

| **Group** | **Mean Satisfaction Score** | **p-value (T-test test; versus placebo)** |
| --- | --- | --- |
| EZC Pak | 0.76 | 0.012 |
| EZC Pak+D | 0.30 | 0.893 |
| EZC Pak Combined | 0.63 | 0.051 |
| Placebo | 0.26 | N/A |

### Patient Safety Data & Side Effects Analysis

In total, 25 participants reported side effects. The most significant side effect was gastrointestinal (GI) discomfort or nausea. This was reported in 6 patients total in the intervention arms, representing 6.5% of the total participants in EZC Pak Combined. This rate of GI discomfort or nausea side effect was within the normal anticipated range given the known potential side effect of high dose zinc.

Three patients in the placebo group reported loose stool or diarrhea. Rice concentrate can have an osmotic effect, especially if taken without additional food. In all but two cases, only a single side effect was reported. One placebo participant reported nausea as well as muscle aches.

Another placebo participant reported headache and drowsiness (Table 7).

**Table 7.** Side Effects Table

| Side Effect n (%) | Placebo Group<br>(n = 90) | EZC Group<br>(n = 60) | EZC D Group<br>(n = 32) | EZC Combined<br>(n=92) |
| --- | --- | --- | --- | --- |
| GI Discomfort/Nausea | 2 (2.2) | 5 (8.3) | 1 (3.1) | 6 (6.5) |
| Vomiting | 0 | 1 (1.7) | 0 | 1 (1.1) |
| Loose Stool/Diarrhea | 3 (3.3) | 1 (1.7) | 0 | 1 (1.1) |
| Dizziness | 0 | 1 (1.7) | 0 | 1 (1.1) |
| Headache | 1 (1.1) | 0 | 0 | 0 |
| Drowsiness | 1 (1.1) | 1 (1.7) | 0 | 1 (1.1) |
| Insomnia | 1 (1.1) | 0 | 0 | 0 |
| Mouth Dryness | 1 (1.1) | 0 | 0 | 0 |
| Nasal Dryness | 0 | 1 (1.7) | 0 | 1 (1.1) |
| Facial Flushing | 0 | 0 | 1 (3.1) | 1 (1.1) |
| Itching | 1 (1.1) | 0 | 0 | 0 |
| Muscle Ache | 2 (2.2) | 0 | 0 | 0 |

## Discussion

The results of this study of EZC Pak’s potential impact on outpatient URI management yield three intriguing results. One, initiating use of EZC Pak as the first step in patients with non-specific URI symptoms has the potential to reduce the duration of illness. The findings support the use of EZC Pak as a viable alternative to patient request for antibiotics when none are deemed clinically necessary at the time of initial clinical presentation. The benefit of EZC Pak on illness duration is likely highest in viral URI. Whether there is a synergistic benefit to using EZC Pak in combination with an antibiotic when a bacterial URI is suspected can be investigated further in future studies.

Secondly, there is a clinically meaningful reduction in symptom severity during URI when using EZC Pak. This lends clinical management to consider EZC Pak alone or in combination with adjuvant treatments that can potentially provide a synergistic reduction in symptom severity during URI (eg, nasal rinse, steam inhalation, or symptom specific relieving medications).

While epidemiologic data suggests patients with low serum vitamin D levels have a higher incidence of URIs^30^, similar to the outcomes of other recent vitamin D trials, the addition of vitamin D during the acute phase of illness in this study neither benefited nor harmed outcomes with respect to illness duration or symptom severity. As such, the decision to replete vitamin D in the acute phase of URI is an individualized decision left to the patient and their clinician.

The patient satisfaction score of EZC Pak compared favorably versus placebo. This highlights an important potential role EZC Pak can play in maintaining or enhancing patient satisfaction in the clinical management of viral URI. This is particularly important given the predominantly self-limited nature of outpatient cases.

EZC Pak can play a critical role in improving outpatient URI management and antibiotic stewardship. Future study can expand the sample size of this initial study to further evaluate these initial key findings.

## Data Availability

All data produced in the present work are contained in the manuscript.

## Notes

### Competing Interest Statement

This clinical trial was carried out by Citrus Labs with funding support from PPC Pharmaceuticals. Aditya Radhakrishnan received a stipend from PPC Pharmaceuticals for his participation in the study. Sarath Malepati is a shareholder in PPC Pharmaceuticals. Stephanie Spencer has no relevant financial disclosures. Naveena Yanamala has no relevant financial disclosures.

### Clinical Trial

NCT04943575

### Author Declarations

Argus Independent Review Board gave ethical approval for this work.

## References

1) O’Neill, J. (2016, May 19). Tackling drug-resistant infections globally: Final report and recommendations. Jim O’Neill. Retrieved from https://apo.org.au/node/63983

2) World Health Organization. (n.d.). Antimicrobial resistance. World Health Organization. Retrieved from http://www.who.int/news-room/fact-sheets/detail/antimicrobial-resistance

3) Sanchez, J. D. (2015, August 4). Paho/WHO: Antimicrobial resistance. Pan American Health Organization / World Health Organization. Retrieved from https://www3.paho.org/hq/index.php?option=com_content&view=article&id=11129%3Aamr-antimicrobial-resistance-intro&Itemid=41534&lang=en#gsc.tab=0

4) Heikkinen, T., & Järvinen, A. (2003). The common cold. The Lancet, 361(9351), 51–59. https://doi.org/10.1016/s0140-6736(03)12162-9

5) Russell CD; Fairfield CJ; Drake TM; Turtle L; Seaton RA; Wootton DG; Sigfrid L; Harrison EM; Docherty AB; de Silva TI; Egan C; Pius R; Hardwick HE; Merson L; Girvan M; Dunning J; Nguyen-Van-Tam JS; Openshaw PJM; Baillie JK; Semple MG; Ho A; ; (n.d.). Co-infections, secondary infections, and antimicrobial use in patients hospitalised with covid-19 during the first pandemic wave from the ISARIC WHO CCP-UK study: A Multicentre, prospective cohort study. The Lancet. Microbe. Retrieved September 22, 2022, from https://pubmed.ncbi.nlm.nih.gov/34100002/

6) Langford, B. J., So, M., Raybardhan, S., Leung, V., Westwood, D., MacFadden, D. R., Soucy, J.-P. R., & Daneman, N. (2020). Bacterial co-infection and secondary infection in patients with COVID-19: A living rapid review and meta-analysis. Clinical Microbiology and Infection, 26(12), 1622–1629. https://doi.org/10.1016/j.cmi.2020.07.016

7) Hu, Y., Lewandowski, E. M., Tan, H., Morgan, R. T., Zhang, X., Jacobs, L. M. C., Butler, S. G., Mongora, M. V., Choy, J., Chen, Y., & Wang, J. (2022, January 1). Naturally occurring mutations of SARS-COV-2 main protease confer drug resistance to Nirmatrelvir. bioRxiv. Retrieved September 22, 2022, from https://www.biorxiv.org/content/10.1101/2022.06.28.497978v1,

8) Sedova, M., Jaroszewski, L., Iyer, M., & Godzik, A. (2022, January 1). Monitoring for SARS-COV-2 drug resistance mutations in broad viral populations. bioRxiv. Retrieved September 22, 2022, from https://www.biorxiv.org/content/10.1101/2022.05.27.493798v1

9) Pfizer reports additional data on paxlovid™ supporting upcoming New Drug Application Submission to U.S. FDA. Pfizer. (n.d.). Retrieved September 22, 2022, from https://www.pfizer.com/news/press-release/press-release-detail/pfizer-reports-additional-data-paxlovidtm-supporting

10) Melchart, D., Linde, K., Worku, F., Bauer, R., & Wagner, H. (1994). Immunomodulation with echinacea — a systematic review of controlled clinical trials. Phytomedicine, 1(3), 245–254. https://doi.org/10.1016/s0944-7113(11)80072-3

11) Barnes, J., Anderson, L. A., Gibbons, S., & Phillipson, J. D. (2005). echinacea species (echinacea angustifolia (DC.) hell., echinacea pallida (Nutt.) Nutt., echinacea purpurea (L.) moench): A review of their chemistry, pharmacology and clinical properties. Journal of Pharmacy and Pharmacology, 57(8), 929–954. ttps://doi.org/10.1211/0022357056127

12) Barrett, B. (2003). Medicinal properties of echinacea: A critical review. Phytomedicine, 10(1), 66–86. https://doi.org/10.1078/094471103321648692

13) Gupta, M., Sharma, D., Sharma, A., Kumari, V., & Goshain, O. P. (n.d.). A Review on Purple Cone Flower (Echinacea purpurea L. Moench). Journal of Pharmacy Research.

14) Mahady, G. B., Qato, D. M., Gyllenhaal, C., Chadwick, L., & Fong, H. H. S. (2001). Echinacea: Recommendations for its use in prophylaxis and treatment of respiratory tract infections. Nutrition in Clinical Care, 4(4), 199–208. https://doi.org/10.1046/j.1523-5408.2001.00143.x

15) Melchart, D., Linde, K., Fischer, P., & Kaesmayr, J. (1999). Echinacea for preventing and treating the common cold. Cochrane Database of Systematic Reviews. https://doi.org/10.1002/14651858.cd000530

16) Shah, S. A., Sander, S., White, C. M., Rinaldi, M., & Coleman, C. I. (2007). Evaluation of echinacea for the prevention and treatment of the common cold: A meta-analysis. The Lancet Infectious Diseases, 7(7), 473–480. https://doi.org/10.1016/s1473-3099(07)70160-3

17) Woelkart, K., Linde, K., & Bauer, R. (2008). Echinacea for preventing and treating the common cold. Planta Medica, 74(6), 633–637. https://doi.org/10.1055/s-2007-993766

18) Petrus, E. J., Lawson, K. A., Bucci, L. R., & Blum, K. (1998). Randomized, double-masked, placebo-controlled clinical study of the effectiveness of zinc acetate lozenges on common cold symptoms in allergy-tested subjects. Current Therapeutic Research, 59(9), 595–607. https://doi.org/10.1016/s0011-393x(98)85058-3

19) Prasad, A. S., Beck, F. W. J., Bao, B., Snell, D., & Fitzgerald, J. T. (2008). Duration and severity of symptoms and levels of plasma interleukin-1 receptor antagonist, soluble tumor necrosis factor receptor, and adhesion molecules in patients with common cold treated with zinc acetate. The Journal of Infectious Diseases, 197(6), 795–802. https://doi.org/10.1086/528803

20) J Eby, G. A. (1997). Zinc ion availability--the determinant of efficacy in zinc lozenge treatment of common colds. Journal of Antimicrobial Chemotherapy, 40(4), 483–493. https://doi.org/10.1093/oxfordjournals.jac.a020864

21) Hemilä, H., Petrus, E. J., Fitzgerald, J. T., & Prasad, A. (2016). Zinc acetate lozenges for treating the common cold: An individual patient data meta-analysis. British Journal of Clinical Pharmacology, 82(5), 1393–1398. https://doi.org/10.1111/bcp.13057

22) Hemilä, H. (1996). Vitamin C and common cold incidence: A review of studies with subjects under heavy physical stress. International Journal of Sports Medicine, 17(05), 379–383. https://doi.org/10.1055/s-2007-972864

23) Hemilä, H. (1997). Vitamin C supplementation and the common cold--was Linus Pauling right or wrong? Int J Vitam Nutr Res. Cochrane Database of Systematic Reviews. (2013). https://doi.org/10.1002/14651858

24) Murdoch, D. R., Slow, S., Chambers, S. T., Jennings, L. C., Stewart, A. W., Priest, P. C., Florkowski, C. M., Livesey, J. H., Camargo, C. A., & Scragg, R. (2012). Effect of vitamin D3supplementation on upper respiratory tract infections in healthy adults. JAMA, 308(13), 1333. https://doi.org/10.1001/jama.2012.12505

25) Jolliffe, D. A., Holt, H., Greenig, M., Talaei, M., Perdek, N., Pfeffer, P., Vivaldi, G., Maltby, S., Symons, J., Barlow, N. L., Normandale, A., Garcha, R., Richter, A. G., Faustini, S. E., Orton, C., Ford, D., Lyons, R. A., Davies, G. A., Kee, F., … Martineau, A. R. (2022). Effect of a test-and-treat approach to vitamin D supplementation on risk of all cause acute respiratory tract infection and covid-19: Phase 3 randomised controlled trial (CORONAVIT). BMJ. https://doi.org/10.1136/bmj-2022-071230

26) Laaksi, I., Ruohola, J. P., Mattila, V., Auvinen, A., Ylikomi, T., & Pihlajamäki, H. (2010). Vitamin D supplementation for the prevention of acute respiratory tract infection: A randomized, double□blinded trial among Young Finnish men. The Journal of Infectious Diseases, 202(5), 809–814. https://doi.org/10.1086/654881

27) Charan, J., Goyal, J. P., Saxena, D., & Yadav, P. (2012). Vitamin D for prevention of respiratory tract infections: A systematic review and meta-analysis. Journal of Pharmacology and Pharmacotherapeutics, 3(4), 300–303. https://doi.org/10.4103/0976-500x.103685

28) Jolliffe, D. A., Camargo, C. A., Sluyter, J. D., Aglipay, M., Aloia, J. F., Ganmaa, D., Bergman, P., Bischoff-Ferrari, H. A., Borzutzky, A., Damsgaard, C. T., Dubnov-Raz, G., Esposito, S., Gilham, C., Ginde, A. A., Golan-Tripto, I., Goodall, E. C., Grant, C. C., Griffiths, C. J., Hibbs, A. M., … Martineau, A. R. (2021). Vitamin D supplementation to prevent acute respiratory infections: A systematic review and meta-analysis of aggregate data from randomised controlled trials. The Lancet Diabetes & Endocrinology, 9(5), 276–292. https://doi.org/10.1016/s2213-8587(21)00051-6

29) Early high-dose vitamin D3 for critically ill, vitamin D–deficient patients. (2019). New England Journal of Medicine, 381(26), 2529–2540. https://doi.org/10.1056/nejmoa1911124

30) Ogunwole, S. U. (2022, March 25). Population under age 18 declined last decade. Census.gov. Retrieved 2022, from https://www.census.gov/library/stories/2021/08/united-states-adult-population-grew-faster-than-nations-total-population-from-2010-to-2020.html

